# Antibody Response after First-dose of ChAdOx1-nCOV (Covishield™^®^) and BBV-152 (Covaxin™^®^) amongst Health Care Workers in India: Preliminary Results of Cross-sectional Coronavirus Vaccine-induced Antibody Titre (COVAT) study

**DOI:** 10.1101/2021.04.07.21255078

**Authors:** Awadhesh Kumar Singh, Sanjeev Ratnakar Phatak, Nagendra Kumar Singh, Arvind Gupta, Arvind Sharma, Kingshuk Bhattacharjee, Ritu Singh

## Abstract

**Background:** Two vaccines are currently being administered in India to prevent the spread of severe acute respiratory syndrome coronavirus 2 (SARS-CoV-2). We assessed the humoral immune response after the first dose of two vaccines ChAdOx1-nCOV (Covishield™) and BBV-152 (Covaxin™) in Indian health care workers (HCW).

**Methods:** This ongoing, Pan-India, Cross-sectional, Coronavirus Vaccine-induced Antibody Titre (COVAT) study is being conducted amongst HCW, with or without past history of SARS-CoV-2 infection. SARS-CoV-2 anti-spike binding antibody is being assessed quantitatively at four timepoints between 21 days or more after the first dose to 6 months after the second dose. Primary aim is to analyze antibody response following each dose of both vaccines and its correlation to age, sex, body mass index (BMI) and comorbidities. Here we report the preliminary results of anti-spike antibody response after the first dose.

**Results:** Amongst the 552 HCW (325 Male, 227 Female), 456 and 96 received first dose of Covishield and Covaxin respectively. Overall, 79.3% showed seropositivity after the first dose. Responder rate and median (IQR) rise in anti-spike antibody was significantly higher in Covishield vs. Covaxin recipient (86.8 vs. 43.8%; 61.5 vs. 6 AU/ml; both p<0.001). This difference persisted in propensity-matched (age, sex and BMI) analysis in 172 subjects. No difference was observed with age, gender and BMI. History of hypertension had lower responder rate (65.7 vs. 82.3%, p=0.001). Covishield recipient had more adverse event vs. Covaxin arm (46.7 vs. 31.2%, p=0.006). Presence of comorbidities, past SARS-CoV-2 infection and vaccine types used were independent predictors for seropositivity after the first dose, in multiple logistic regression analysis.

**Conclusions:** While both vaccines elicited immune response, seropositivity rates to anti-spike antibody were significantly higher in Covishield recipient compared to Covaxin after the first dose. Ongoing COVAT study will further enlighten the immune response between two vaccines after the second dose.

**Highlights:**

1. This study evaluated the humoral antibody response of two SARS-CoV-2 vaccines Covishield™ and Covaxin™ in Indian health-care workers.
2. Both vaccines showed seropositivity to anti-spike antibody, 21 days or more after the first dose.
3. Responder rates were higher in Covishield recipient compared to Covaxin in propensity-matched cohorts.
4. Past SARS-CoV-2 infection, presence of comorbidities and vaccine type received were independent predictors of antibody response after the first dose.

## 1. Introduction

The pandemic of Coronavirus disease 2019 (COVID-19) caused by Severe Acute Respiratory Syndrome Coronavirus 2 (SARS-CoV-2) has already affected more than 127 million people and caused more than 2.7 million deaths worldwide, as of March 30, 2021 [1]. To contain this brunt, several novel vaccines have recently received an Emergency Use Authorization (EUA) by the U.S.A Food Drug Administration (FDA), European Medicine Agency (EMA), U.K. Medicines and Healthcare Products Regulatory Agency (MHRA) and Indian Central Drugs Standard Control Organization (CDSCO) as well as Drugs Controller General of India (DCGI). After receiving EUA, these vaccines are currently being administered to health-care workers, front-line workers, elderly and at-risk individuals, including people with comorbidities, in orderly fashion. An estimate by World Health Organization on March 30, 2021 reports more than 520 million vaccine doses have been so far administered [1]. The Indian national vaccination program started from January 16, 2021 after the approval of two candidate vaccines namely Covishield™ (ChAdOx1-nCOV or AZD1222, acquired from Oxford University and AstraZeneca, manufactured by Serum Institute of India, Pune) and Covaxin™ (BBV-152, manufactured by Bharat Biotech, Hyderabad in collaboration with Indian Council of Medical Research [ICMR], India). While, early data from available phase 3 clinical trials do suggest that these two vaccines are safe and effective [2-4], there is still a paucity of information as to how much and how long, these novel vaccines can elicit an immune response, both at cellular as well as the molecular level. Notably, while, both vaccines Covishield and Covaxin have been approved for the two dosage at the interval of 4-weeks in India, a gap of 4- 6 weeks initially and up to 8-weeks recently for the second dose has been allowed for the Covishield by the CDSCO and DCGI.

It is well-known that a humoral immune response to an acquired infection or vaccination, results in two major immune changes in human. The antibodies produced by the antibody secreting cells (ASC) provide a rapid, protective immunity and, the generation of long-lived memory B cells become capable of mounting recall responses whenever re-exposed. Interestingly, following re-exposure when circulating antibodies fails to confer protection, the memory B cells drive the recall response by producing new antibodies either through the formation of new ASC or re-initiating germinal center reactions to generate new, high-affinity B cell clones via an additional round of somatic hypermutation [5]. Available evidence suggests that, these immunological memories in the form of antibodies and memory B cells are durable for over 8 months post-SARS-CoV-2 infection [6, 7] although the exact duration of immune protection after the infection or vaccination is currently unknown. A robust immunogenic response until 3-month after the second dose of Moderna mRNA-1273 vaccine has been recently reported [8]. While the antibody kinetics, its association with severity and correlation to protection has been extensively reviewed with other vaccines [9, 10], the antibody kinetics after vaccination with Covishield and Covaxin is less well known. Phase 1/2 studies that have evaluated the kinetics of both short-term binding (against all epitopes such as spike protein, receptor-binding domain and nucleocapsid protein) and neutralizing antibodies have demonstrated a significant rapid increase with both Covishield and Covaxin in vaccinated SARS-CoV-2 naїve as well as SARS-CoV-2 recovered individuals [11-13]. However, there is still limited knowledge about antibody repertoire over the time and its variation and correlation to age, sex, body mass index (BMI), and in presence of comorbidities including its duration and treatment, especially in Indians. Moreover, it is also not yet known whether the antibody kinetics differ between the two vaccines approved in India, after the first and second dose, and thereafter over time. Furthermore, the antibody repertoire between the two vaccines in SARS-CoV-2 naїve and SARS-CoV-2 recovered individuals is not entirely known in Indians.

These findings prompted us to conduct a cross-sectional study to evaluate a longitudinal humoral response on the kinetics of anti-spike binding antibody formation, following both the first and second dose of Covishield and Covaxin, until 6-months after its completion. This study has involved participants with both SARS-CoV-2 naїve and SARS-CoV-2 recovered individuals. In this preliminary analysis, we report the binding anti-spike antibody kinetics after the first dose (day 21 or more until the second dose of vaccine) of two vaccine from ongoing Coronavirus Vaccine-induced Antibody Titre (COVAT) study.

## 2. Methods

### 2.1 Study design and participants

This report followed the Strengthening the Reporting of Observational Studies in Epidemiology (STROBE) reporting guideline for cross-sectional studies [14]. COVAT study is an ongoing, pan-India, cross-sectional study that was approved by the ethical committee of Thakershy Charitable Trust, Ahmedabad, Gujrat, India. All adult health care workers of more than 18 years of age who received the first dose of vaccine were eligible to participate in this study including those who had recovered from the COVID-19 in the recent past (> 6 weeks before the first dose). Individuals with current confirmed SARS-CoV-2 infection and those diagnosed within 6-weeks were excluded from the study. Written informed consent were taken from all the participants who participated in this study, voluntarily.

### 2.2 Measurements

Clinical data was collected from all eligible participants including age, sex, blood groups, body mass index (BMI), past history of confirmed SARS-CoV-2 infection, presence of comorbidities such as diabetes mellitus (type 1 [T1DM] and type 2 [T2DM]), hypertension (HTN), dyslipidemia, presence of ischemic heart disease (IHD), chronic kidney disease (CKD) and cancer, including its duration and treatment received. In addition, we have also collected the data regarding any adverse events post-vaccination and subsequent SARS-CoV-2 infection after the first or second dose of vaccine and data collection will be continued until 6-month after the second dose.

Anti-spike antibody titre would be measured at four time-points: day 21 after the first dose until the day before the second dose; day 21-28 of second dose, day 83-97 (3-month) and day 173-187 (6-month) after the second dose. At first, blood samples (5 ml) were collected from eligible health-care workers (all of whom are doctors by profession) day-21 onwards, until the day before the second dose. All samples were collected as either serum or plasma using EDTA vials from each participant and analyzed at Central laboratory of Neuberg, Supratech at Ahmedabad, Gujarat, India. The IgG antibodies to SARS-CoV-2 directed against the spike protein (S-antigen, both S1 and S2 protein) were assayed with LIASON^®^ S1/S2 quantitative antibody detection kit (DiaSorin Saluggia, Italy) using indirect chemiluminescence immunoassay (CLIA) as per manufacturer’s protocol. The sensitivity of the above assay is 97.4% and specificity is 98.9% at 15 days post-diagnosis for laboratory serological [12]. Testing of assay-specific calibrators allows the detected Relative light unit (RLU) to adjust the assigned master curve. The analyzer automatically calculates SARS-CoV-2 S1/S2 IgG antibody concentrations as arbitrary units (AU/mL) and grades the results. Antibody levels ≥ 15.0 arbitrary unit (AU)/ml were considered as sero-positive or responders, while antibody level <15 AU/ml are considered as seronegative or non-responders, as per manufacturer’s kit. The lower and upper limit of this quantitative spike antibody kit is 3.8 and 400 AU/ml respectively, as per manufacture’s brochure.

### 2.3 Statistical analysis

Descriptive and inferential statistical analysis has been carried out for the present study. Normality of the data was assessed by Shapiro-Wilk test and visually by QQ plot for Covishield and Covaxin subgroups. Data on continuous scale were presented as Median (Interquartile range, IQR) and categorical data were presented as number (%). A two-sided P value of < 0.05 was considered as statistically significant. Chi-square test was used to find the significance of study parameters on categorical scale between two or more groups. Mann-Whitney test was utilized to assess two non-parametric groups and Kruskal-Wallis test was used to compare the differences among two or multiple data group for data on continuous scale. To compare the antibody kinetics between two vaccines, we also carried out a propensity-matched comparison of two groups of participants. A propensity score was generated taking into consideration age, sex and BMI of the individual participants. Participants having similar scores were matched and two groups were compared accordingly. Moreover, multiple logistic regression analysis was also conducted to find out whether any independent factors were associated with a blunted response to vaccine in anti-spike antibody generation following first dose of vaccination. Entire statistical analysis was carried out with Statistical Package for Social Sciences (SPSS Complex Samples) Software Version 22.0 for windows, SPSS Inc., Chicago, IL, USA, with Microsoft Word and Excel being used to generate graphs and tables.

## 3. Results

Five hundred fifty-two participants who received first dose of either of the two vaccines had complete set of data including anti-spike antibody. The mean age of the participants was 44.8 ± 13.2 years, with 58.9% males (325/552) and 41.1% females (227/552). Out of 552 participants, 464 were aged ≤ 60 years and 88 were of age > 60 years. While 23.7% (131/552) had one or more comorbidities (10.3% T2DM, 17.9% HTN, 4.7% dyslipidemia and 2.3% IHD), we had no participant having T1DM, CKD or cancer in our study cohort. Amongst participants, 10.9% (60/552) had past history of confirmed SARS-CoV-2 who took the first dose of either of the vaccine.

### 3.1 SARS-COV-2 spike antibody positivity rates after first dose of two vaccine

A total of 552 (325 male, 227 female) participants’ data was analyzed. Out of these 456 and 96 received first dose of Covishield and Covaxin respectively. Overall, 79.3% (438/552) had seropositivity and were responders (defined as anti-spike antibody titre ≥ 15AU/ml measured at day 21 to day before second dose) for anti-spike antibody. Intriguingly, the responder rate was significantly higher in Covishield vs. Covaxin recipient (86.8 vs. 43.8% respectively, p<0.001). No association was found between age and antibody response in the overall and Covaxin cohort, however a greater responder rate was elicited in the age ≤ 60 years for the Covishield cohort when compared to age > 60 years (88.3% vs. 79.2% respectively, p=0.036). Presence of any comorbidities was associated with a lower responder rate, compared to those without (72.5% vs. 81.5% respectively, p=0.027). This finding appears to be driven by the Covishield arm having a significantly lower responder rate in presence of comorbidities compared to those without (78.0% vs. 89.9%, p=0.001). Amongst the captured co-morbidities, only history of HTN was associated with a significantly lower response rate compared to those without (65.7% vs. 82.3%, p=0.001), and it was mainly driven by lower responder rate in Covishield arm (72.7% vs 89.7%, p <0.001). Notably, past history of SARS-CoV-2 infection resulted in a significantly greater antibody responder rate compared to SARS-CoV-2 naїve individuals in overall cohort (96.7% vs. 77.2%, p<0.001), irrespective of the types of vaccine received. No differential antibody responder rate was observed with regard to gender, body mass index (BMI), blood groups, presence of T2DM, dyslipidemia and IHD including its duration and management strategies. (Table 1).

**Table 1:** **Responders to anti-spike antibody after the first dose of either vaccine, at day 21 or more but before the second dose**

| Characteristics | Overall, N= 552 |  | Covishield, N= 456 |  | Covaxin, N= 96 |  |
| --- | --- | --- | --- | --- | --- | --- |
|  | Responder Rate | P | Responder Rate | P | Responder Rate | P |
| Age ≤ 60 years, n (%) | 372/464 (80.2) | 0.272 | 339/384 (88.3) | 0.036 | 33/80 (41.2) | 0.270 |
| Age > 60 years, n (%) | 66/88 (74.4) |  | 57/72 (79.2) |  | 9/16 (56.2) |  |
| Male, n (%) | 251/325 (77.2) | 0.142 | 228/268 (85.1) | 0.182 | 23/57 (40.4) | 0.417 |
| Female, n (%) | 187/227 (82.4) |  | 168/188 (89.4) |  | 19/39 (48.7) |  |
| BMI < 25 kg/m<sup>2</sup> | 301/379 (79.4) | 0.731 | 273/309 (88.3) | 0.375 | 27/70 (38.6) | 0.150 |
| BMI 25-29.9 kg/m<sup>2</sup> | 85/106 (80.2) |  | 74/89 (83.1) |  | 11/17 (64.7) |  |
| BMI ≥ 30 kg/m<sup>2</sup> | 57/67 (85.1) |  | 49/58 (84.5) |  | 4/9 (44.4) |  |
| Any Co-morbidities, n (%) | 95/131 (72.5) | 0.027 | 92/118 (78) | 0.001 | 3/13 (23.1) | 0.092 |
| No Co-morbidities, n (%) | 343/421 (81.5) |  | 304/338 (89.9) |  | 39/83 (47) |  |
| T2DM, n (%) | 42/57 (73.7) | 0.265 | 39/49 (79.6) | 0.112 | 3/8 (37.5) | 0.710 |
| No T2DM, n (%) | 396/495 (80) |  | 357/407 (87.7) |  | 39/88 (44.3) |  |
| T2DM Duration < 5 years, n (%) | 9/11 (81.8) | 0.751 | 8/10 (80) | 0.994 | 1/1 (100) | 0.214 |
| T2DM Duration 5-10 years, n (%) | 21/31 (67.74) |  | 20/26 (76.92) |  | 1/5 (20) |  |
| T2DM Duration > 10 years, n (%) | 11/15 (73.3) |  | 11/14 (78.6) |  | 0/1 (0) |  |
| T2DM-Optimum control, n (%) | 40/55 (72.7) | 0.390 | 38/48 (79.2) | 0.609 | 2/7 (28.6) | 0.168 |
| T2DM-No optimum control, n (%) | 0/2 (0) |  | 1/1 (100) |  | 1/1 (100) |  |
| *T2DM Management* |  |  |  |  |  |  |
| Monotherapy, n (%) | 14/16 (87.5) | 0.287 | 12/13 (92.3) | 0.414 | 2/3 (66.7) | 0.155 |
| Combination therapy, n (%) | 24/35 (68.6) |  | 24/32 (75) |  | 0/3 (0) |  |
| Insulin, n (%) | 1/1 (100) |  | 1/1 (100) |  | 0 (0) |  |
| No medication, n (%) | 3/5 (60) |  | 3/4 (75) |  | 0/1 (0) |  |
| HTN, n (%) | 65/99 (65.7) | 0.001 | 56/77 (72.7) | <0.001 | 13/22 (55.4) | 0.760 |
| No HTN, n (%) | 373/453 (82.3) |  | 340/379 (89.7) |  | 41/74 (88.9) |  |
| HTN Duration < 5 years, n (%) | 17/29 (58.6) | 0.549 | 17/26 (65.4) | 0.414 | 0/3 (0) | 0.482 |
| HTN Duration 5-10 years, n (%) | 33/48 (68.8) |  | 25/31 (80.6) |  | 8/17 (47.1) |  |
| HTN Duration > 10 years, n (%) | 15/22 (68.2) |  | 14/20 (70) |  | 1/2 (50) |  |
| *HTN Management* |  |  |  |  |  |  |
| RAAS Blockers, n (%) | 28/41 (68.3) | 0.944 | 27/37 (73) | 0.928 | 1/4 (25) | 0.704 |
| CCBs, n (%) | 14/22 (63.6) |  | 14/20 (70) |  | 0/2 (0) |  |
| Combined, n (%) | 14/21 (66.7) |  | 14/19 (73.7) |  | 0/2 (0) |  |
| No Medicine, n (%) | 1/2 (50) |  | 1/1 (100) |  | 0/1 (0) |  |
| Dyslipidemia, n (%) | 23/26 (88.5) | 0.437 | 23/26 (88.5) | 0.897 | 0 | NA |
| No Dyslipidemia, n (%) | 415/526 (78.89) |  | 373/430 (86.7) |  | 42/96 (43.8) |  |
| IHD, n (%) | 11/13 (84.6) | 0.635 | 11/13 (84.6) | 0.810 | 0 |  |
| No IHD, n (%) | 427/539 (79.2) |  | 385/443 (86.9) |  | 42/96 (43.8) |  |
| Past H/o COVID-19, n (%) | 58/60 (96.7) | <0.001 | 55/57 (96.5) | 0.023 | 3/3 (100) | 0.046 |
| No Past H/o COVID-19, n (%) | 380/492 (77.2) |  | 341/399 (85.5) |  | 39/93 (41.9) |  |
| Covishield, n (%) | 396/456 (86.8) | <0.001 |  |  |  |  |
| Covaxin, n (%) | 42/96 (43.8) |  |  |  |  |  |
| p<0.05 considered as statistically significant, p computed by chi-square test, BMI- body mass index, T2DM- type 2 diabetes mellitus, HTN- hypertension, RAAS- Renin angiotensin aldosterone system, CCB- calcium channel blocker, IHD- ischemic heart disease, COVID-19- coronavirus disease 2019 |  |  |  |  |  |  |

### 3.2 Assessment of SARS-CoV-2 spike antibody quantity after first dose of two vaccine

In line with the antibody responder rate, the Covishield arm elicited a differential quantitative antibody titer response. The median (IQR) rise in anti-spike antibody (61.5 vs. 6 AU/ml, p<0.001) was significantly higher in Covishield vs. Covaxin recipient. Higher median values of anti-spike antibody in age ≤ 60 years was also observed compared to the age >60 years, which was highly significant in Covishield arm (65.0 vs. 42.5 AU/ml respectively, p=0.01). In the overall cohort, the females had a significantly higher median antibody titre compared to the males (67.0 vs. 47.0 AU/ml, p=0.006), primarily driven by Covishield arm. Presence of comorbidities also resulted in lower median antibody titre in the overall cohort compared to those without, however this was significantly noticeable in Covishield arm (54.0 vs. 64.5 AU/ml, p=0.03). Amongst comorbidities, history of HTN was associated with a significantly lower median antibody titre compared to those without (33.0 vs. 60.0 AU/ml, p=0.001), particularly in the Covishield arm. Indeed, past history of infection of SARS-CoV-2 elicited a significantly greater median antibody titre in the overall cohort, compared to SARS-CoV-2 naїve (400.0 vs. 48.0 AU/ml, p <0.001), irrespective of types of vaccine received. Notably, Covishield arm had a significantly higher median antibody titer compared to Covaxin arm (61.5 vs. 6.0 AU/ml respectively, p <0.001). Table 2 summarizes the results of antibody titre across all groups. Box and whisker plot in figure 1 depicts the antibody titre for different study parameters that were significantly different after the first dose of vaccine. With regard to timing of sampling after the first dose, there was no significant difference in median antibody titre when measured between day 21-28 or 29-36 or >37 after the first dose of vaccine (Figure 2, supplementary table 1).

**Table 2:** **Median (interquartile range) anti-spike antibody after the first dose of either vaccine, at day 21 or more but before the second dose**

| Characteristics | Overall, N= 552 |  | Covishield, N= 456 |  | Covaxin, N= 96 |  |
| --- | --- | --- | --- | --- | --- | --- |
|  | Antibody Titer,<br>Median (IQR)<br>(in AU/ml) | P | Antibody Titer,<br>Median (IQR)<br>(in AU/ml) | P | Antibody Titer,<br>Median (IQR)<br>(in AU/ml) | P |
| Age ≤ 60 years | 60 (23-117.5) | 0.055 | 65 (34-128.75) | 0.014 | 19.5 (4-98) | 0.605 |
| Age > 60 years | 37 (15.25-101.25) |  | 42.5 (18.75-101.25) |  | 5.5 (4-99.7) |  |
| Male | 47 (17.5-105) | 0.006 | 52.5 (26-118.25) | 0.004 | 5 (4-95) | 0.219 |
| Female | 67 (30 – 122) |  | 70 (41-157.25) |  | 7 (4-110) |  |
| BMI < 25 kg/m<sup>2</sup> | 57 (21-113) | 0.667 | 62 (31.5-118.5) | 0.946 | 5 (4-77.75) | 0.189 |
| BMI 25-29.9 kg/m<sup>2</sup> | 55 (23.3-119.75) |  | 58 (27-123.5) |  | 36 (4-159.5) |  |
| BMI ≥ 30 kg/m<sup>2</sup> | 59 (21-157) |  | 59.5 (28.5-309.75) |  | 7 (4.9 – 113.5) |  |
| Any Co-morbidities | 46 (10-100) | 0.059 | 54 (20.5-110) | 0.032 | 4 (4-12.5) | 0.058 |
| No Co-morbidities | 60 (25-120) |  | 64.5 (35-122.75) |  | 7 (4-106) |  |
| T2DM | 40 (8.5-95) | 0.186 | 47 (21.5 – 152) | 0.246 | 4 (4-20.25) | 0.198 |
| No T2DM | 58 (23-118) |  | 62 (33-120) |  | 6.37 (4-101.5) |  |
| T2DM Duration < 5 year | 42 (21-100) | 0.697 | 43.5 (23-175) | 0.949 | 21 (21-21) | 0.165 |
| T2DM Duration 5-10 year |  |  |  |  |  |  |
| T2DM Duration > 10 year | 45 (6.25-82) |  | 68 (19.5-83) |  | 4 (4-11) |  |
|  | 31 (9-400) |  | 31.5 (18.75-400) |  | 4 (4-4) |  |
| T2DM-Optimum control | 40 (8-90) | 0.752 | 49.5 (21.25-178) | 0.531 | 4 (4-18) | 0.250 |
| T2DM-No optimum control | 58 (23-118) |  | 62 (33-120) |  | 6.37 (4-101.5) |  |
| *T2DM Management* |  |  |  |  |  |  |
| Monotherapy | 51 (21-83.5) |  | 80 (33-92) |  | 18 (4-18) |  |
| Combination therapy | 32 (8-204) | 0.811 | 35 (11.25-293.25) | 0.648 | 4 (4-4) | 0.211 |
| Insulin | 47 (4-241) |  | 64.5 (14.75-320.5) |  | 4 (4-4) |  |
| No medication | 58.5 (23-117.5) |  | 62 (33-120) |  | 6.74 (4-103.5) |  |
| HTN | 33 (8-89) | 0.001 | 40 (13.5-86) | <0.001 | 5.86 (4-110) | 0.808 |
| No HTN | 60 (26-122) |  | 65 (35-157) |  | 6 (4-97) |  |
| HTN Duration < 5 years | 30 (8-73.5) |  | 41 (8.75-81.25) |  | 4 (4-4) |  |
| HTN Duration 5-10 years | 37 (7-118.45) | 0.659 | 52 (21-90) | 0.591 | 7 (4-126.5) | 0.270 |
| HTN Duration > 10 years | 24 (9-92.25) |  | 24 (11.25-87) |  | 54.5 (4-54.5) |  |
| *HTN Management* |  |  |  |  |  |  |
| RAAS Blockers | 33 (9-33) |  | 40 (13.5-83.5) |  | 5.5 (4-80.5) |  |
| CCBs | 23 (6-71.5) | 0.946 | 25 (8-82.5) | 0.650 | 4 (4-4) | 0.319 |
| Combined | 38 (9-132) |  | 46 (15-174) |  | 4 (4-4) |  |
| No Medicine | 32.5 (4-32.5) |  | 61 (61-61) |  | 4 (4-102) |  |
| Dyslipidemia | 52.5 (27-110) | 0.671 | 52.5 (27-110) | 0.431 | NA | NA |
| No Dyslipidemia | 58 (21-115) |  | 62 (31.5-123.5) |  | 6 (4-99.7) |  |
| IHD | 38 (22.5-102) | 0.492 | 38 (22.5-102) | 0.202 | NA | NA |
| No IHD | 58 (22-115) |  | 62 (31-121) |  | 6 (4-99.7) |  |
| Past H/o COVID-19 | 400 (294-400) | <0.001 | 400 (298-400) | <0.001 | 260 (214-260) | 0.001 |
| No Past H/o COVID-19 | 48 (18.25-94) |  | 55 (28-94) |  | 5 (4-95) |  |
| Covishield | 61.5 (30-119.5) | <0.001 |  |  |  |  |
| Covaxin | 6 (4-99.7) |  |  |  |  |  |
| p<0.05 considered as statistically significant, p computed by Mann-Whitney test of Kruskal-Wallis test, BMI- body mass index, T2DM- type 2 diabetes mellitus, HTN- hypertension, RAAS- Renin angiotensin aldosterone system, CCB- calcium channel blocker, IHD- ischemic heart disease, COVID-19- coronavirus disease 2019 |  |  |  |  |  |  |

**Figure 1:**
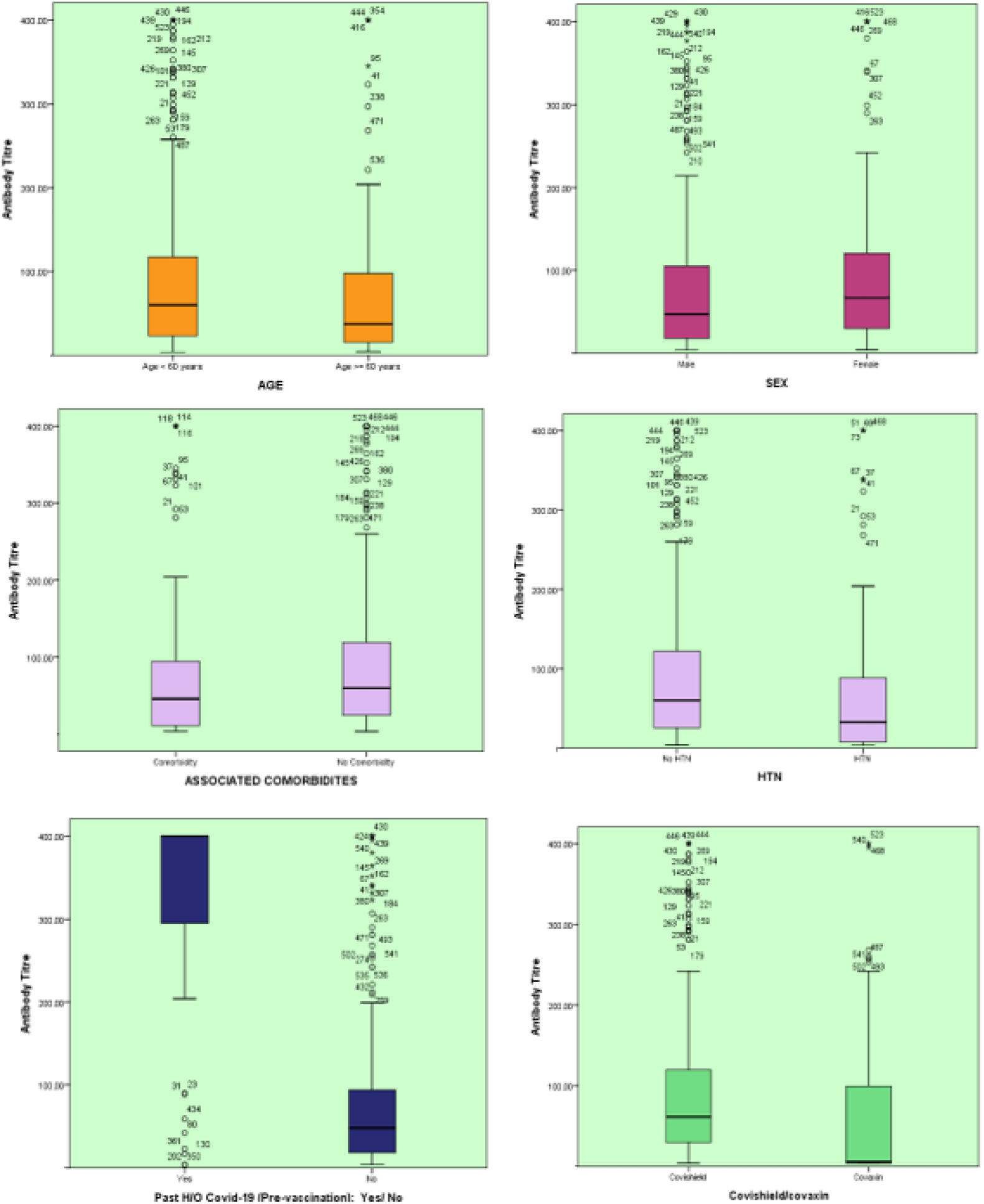
Box and whisker plot demonstrating antibody titer by study parameters.

**Figure 2:**
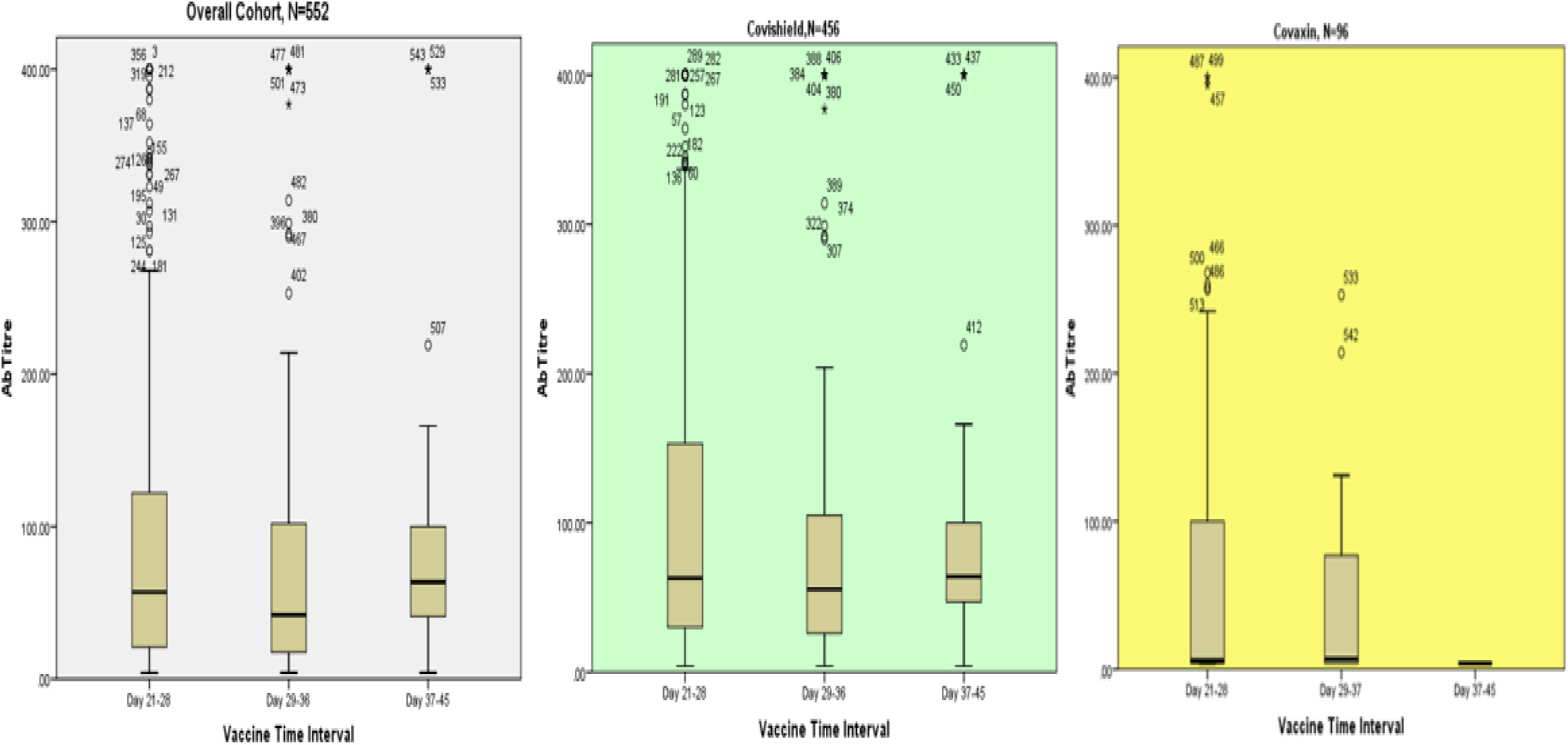
Box and whisker plot demonstrating antibody titer between Day 21-28, 29-36 and 36-45 days after the first dose

### 3.3 Comparison of SARS-CoV-2 spike antibody between Covishield and Covaxin after propensity matched analysis

In the propensity-matched analysis of 172 cohorts (86 participants in each arm) after the adjustment for age, sex and BMI; a past history of SARS-CoV-2 infection and type of vaccine received were found to be associated with a differential antibody responder rate. Responder rates were significantly higher in Covishield arm compared to the Covaxin arm (88.4% vs. 43.0%, p <0.001) in the propensity-matched analysis. Interestingly, in Covishield arm, presence of T2DM was associated with poor antibody response compared to those without T2DM (63.6% vs. 92.0%, p=0.006) while presence of any comorbidities elicited a poor antibody response compared to those without (9.1% vs. 48.0%, p=0.02) in Covaxin arm, in the propensity-matched analysis (Table 3).

**Table 3:** **Responders to anti-spike antibody after the first dose of either vaccine, at day 21 or more but before the second dose - Propensity Score (Age, Sex, BMI) Matched Analysis**

| Characteristics | Overall, N= 172 |  | Covishield, N= 86 |  | Covaxin, N= 86 |  |
| --- | --- | --- | --- | --- | --- | --- |
|  | Responder Rate | P | Responder Rate | P | Responder Rate | P |
| Age ≤ 60 years, n (%) | 98/148 (66.2) | 0.722 | 67/73 (91.2) | 0.019 | 31/75 (41.3) | 0.409 |
| Age > 60 years, n (%) | 15/24 (62.5) |  | 9/13 (69.2) |  | 6/11 (54.5) |  |
| Male, n (%) | 52/81 (64.2) | 0.696 | 36/40 (90) | 0.661 | 16/41 (39) | 0.475 |
| Female, n (%) | 61/91 (67) |  | 40/46 (87) |  | 21/45 (46.7) |  |
| BMI < 25 kg/m<sup>2</sup> | 78/124 (62.9) | 0.402 | 55/62 (88.7) | 0.938 | 23/62 (37.1) | 0.116 |
| BMI 25-29.9 kg/m<sup>2</sup> | 22/29 (75.9) |  | 12/14 (85.7) |  | 10/15 (66.7) |  |
| BMI ≥ 30 kg/m<sup>2</sup> | 13/19 (68.4) |  | 9/10 (90) |  | 4/9 (44.4) |  |
| Any Co-morbidities, n (%) | 19/33 (57.6) | 0.274 | 18/22 (81.8) | 0.266 | 1/11 (9.1) | 0.015 |
| No Co-morbidities, n (%) | 94/139 (67.6) |  | 58/64 (90.6) |  | 36/75 (48) |  |
| T2DM, n (%) | 8/17 (47.1) | 0.088 | 7/11 (63.6) | 0.006 | 1/6 (16.7) | 0.176 |
| No T2DM, n (%) | 105/155 (67.7) |  | 69/75 (92) |  | 36/80 (45) |  |
| T2DM Duration < 5 years, n (%) | 3/4 (75) | 0.114 | 2/3 (66.7) | 0.059 | 1/1 (100) | 0.156 |
| T2DM Duration 5-10 years, n (%) | 4/8 (50) |  | 4/4 (100) |  | 0/4 (0) |  |
| T2DM Duration > 10 years, n (%) | 1/5 (20) |  | 1/4 (25) |  | 0/1 (0) |  |
| T2DM-Optimum control, n (%) | 7/16 (43.8) | 0.121 | 6/10 (60) | 0.052 | 1/6 (16.7) | 0.176 |
| T2DM-No optimum control, n (%) | 1/1 (100) |  | 1/1 (100) |  |  |  |
| *T2DM Management* |  |  |  |  |  |  |
| Monotherapy, n (%) | 4/6 (66.6) |  | 3/3 (100) |  | 1/3 (33.3) |  |
| Combination therapy, n (%) | 3/10 (30) | 0.085 | 3/7 (42.9) | 0.198 | 0/3 (0) | 0.364 |
| Insulin, n (%) | 1/1 (100) |  | 1/1 (100) |  | 0/1 (0) |  |
| No medication, n (%) |  |  |  |  |  |  |
| HTN, n (%) | 93/137 (67.9) | 0.232 | 64/72 (88.9) | 0.735 | 29/65 (44.6) | 0.600 |
| No HTN, n (%) | 20/35 (57.1) |  | 12/14 (85.7) |  | 8/21 (38.1) |  |
| HTN Duration < 5 years, n (%) | 4/8 (50) | 0.645 | 4/5 (80) | 0.634 | 0/3 (0) | 0.362 |
| HTN Duration 5-10 years, n (%) | 13/22 (59.1) |  | 5/5 (100) |  | 8/17 (47.1) |  |
| HTN Duration > 10 years, n (%) | 3/5 (60) |  | 3/4 (75) |  | 1/1 (0) |  |
| *HTN Management* |  |  |  |  |  |  |
| RAAS Blockers, n (%) | 4/8 (50) |  | 4/5 (80) |  | 0/3 |  |
| CCBs, n (%) | 3/6 (50) | 0.749 | 3/4 (75) | 0.788 | 0/2 | 0.155 |
| Combined, n (%) | 4/6 (66.7) |  | 4/4 (100) |  | 0/2 |  |
| No Medicine, n (%) | 1/2 (50) |  | 1/1 (100) |  | 0/1 |  |
| Dyslipidemia, n (%) | 8/10 (80) | 0.768 | 6/6 (100) | 0.102 | 2/4 (50) | 0.864 |
| No Dyslipidemia, n (%) | 107/162 (66.04) |  | 63/80 (78.75) |  | 44/82 (53.69) |  |
| IHD, n (%) | 3/4 (75) | 0.692 | 3/4 (75) | 0.393 | 0/0 | 0.393 |
| No IHD, n (%) | 110/168 (65.5) |  | 73/82 (89) |  | 37/86 (43) |  |
| Past H/o COVID-19, n (%) | 12/12 (100) | 0.009 | 9/9 (100) | 0.250 | 3/3 (100) | 0.042 |
| No Past H/o COVID-19, n (%) | 101/160 (63.1) |  | 67/77 (87) |  | 34/83 (41) |  |
| Covishield, n (%) | 76/86 (88.4) | <0.001 |  |  |  |  |
| Covaxin, n (%) | 37/86 (43) |  |  |  |  |  |
| BMI- body mass index, T2DM- type 2 diabetes mellitus, HTN- hypertension, RAAS- Renin angiotensin aldosterone system, CCB- calcium channel blocker, IHD- ischemic heart disease, COVID-19- coronavirus disease 2019 |  |  |  |  |  |  |

### 3.4 Independent variables associated with responder vs. Non-responder to spike antibody

Multiple logistic regression analysis (to identify the independent predictors for non-responder rate based on development of SARS-CoV-2 Anti-spike antibody) suggests that amongst all the variables analyzed, three variables such as- presence of comorbidities, past history of SARS-CoV-2 infection and vaccine type were independent predictors of antibody response rate. While associated co-morbidities were significantly related to an increase in the non-responder rate (Odds Ratio [OR] 2.73; 95% Confidence Interval [CI], 1.17 - 6.39; p=0.021), previous SARS-CoV-2 infection significantly reduced the non-responder rate by 86% (OR 0.14; 95% CI, 0.03 - 0.61; p=0.009). Notably, recipients of Covishield had a significantly lower non-responder rate by 89% (OR 0.11; 95% CI, 0.06 - 0.19; p<0.001), compared to those who received Covaxin (Table 4).

**Table 4:** **Multiple logistic regression to identify the independent predictors for non-responder rate (based on development of SARS CoV2 Anti-spike antibody):**

| Variables in the Equation |  |  |  |  |  |  |  |  |
| --- | --- | --- | --- | --- | --- | --- | --- | --- |
|  | B | S.E. | Wald | df | P value | Exp(B) | 95% C.I. for EXP(B) |  |
|  |  |  |  |  |  |  | Lower | Upper |
| Age | .019 | .327 | .003 | 1 | .954 | 1.019 | .537 | 1.935 |
| Sex | .281 | .250 | 1.263 | 1 | .261 | 1.324 | .811 | 2.161 |
| BMI | -.022 | .176 | .016 | 1 | .900 | .978 | .692 | 1.382 |
| Associated Comorbidities | 1.004 | .434 | 5.366 | 1 | .021 | 2.730 | 1.167 | 6.386 |
| Type 2 diabetes | .163 | .470 | .120 | 1 | .729 | 1.177 | .468 | 2.958 |
| Hypertension | -.323 | .383 | .714 | 1 | .398 | .724 | .342 | 1.532 |
| Ischemic heart disease | -.236 | .950 | .062 | 1 | .803 | .789 | .123 | 5.084 |
| Dyslipidaemia | .000 | .000 | 2.333 | 1 | .127 | 1.000 | 1.000 | 1.000 |
| Past H/O Covid-19 | -1.947 | .744 | 6.848 | 1 | .009 | .143 | .033 | .613 |
| Covishield vs. Covaxin | -2.206 | .274 | 64.631 | 1 | <0.001 | .110 | .064 | .189 |
| Constant | 6.929 | 1.884 | 13.528 | 1 | .0001 | 1021.857 |  |  |
| a. Variable(s) entered on regression step: Age, Sex, BMI (body mass index), Associated Comorbidities, Type 2 diabetes, Hypertension, Ischemic heart disease, Dyslipidaemia, Past H/O Covid-19, Covishield/Covaxin. |  |  |  |  |  |  |  |  |

### 3.5 Post-vaccination adverse events and post-vaccination SARS-CoV-2 infection

In the overall post-vaccination cohort, the Covishield arm had a higher proportion of patients having any side-effects as compared to Covaxin arm (46.7% vs. 31.2%, p=0.006). Notably, in the propensity-matched cohort, any side-effects post vaccination was also higher in the Covishield compared to the Covaxin arm (50.0 % vs. 30.2%, p=0.008). Until writing of this manuscript, two participants from the Covishield arm reported to have a suspected (RT-PCR negative conducted twice) and confirmed (RT-PCR positive) symptomatic COVID-19 respectively. While both had mild COVID-19 and none required hospitalization; suspected COVID-19 was reported within 3-weeks and confirmed COVID-19 was reported 4-weeks after the second dose of Covishield vaccine.

## 4. Discussion

Summarily, this cross-sectional COVAT study overall reported a 79.3% responder rate after the first dose of both vaccines. There was no significant difference in responder rate with regard to age, sex, BMI, blood group and any comorbidities including its duration and treatment, except that the participants with history of HTN had significantly less responder rate and median antibody titre, compared to those without. Importantly, median titre of antibody was significantly higher in females compared to the males. Past history of SARS-CoV-2 infection had a significantly higher responder rate and median anti-spike antibody titre compared to the SARS-CoV-2 naїve individuals, after the first dose of either of the two vaccines. Any adverse side effects post-vaccination was significantly higher in Covishield recipient compared to the Covaxin; however, these adverse events were mild to moderate in nature. Intriguingly, the responder rate and median rise in anti-spike antibody was significantly higher with Covishield recipient, compared to the Covaxin. Whether this differential finding between two vaccines is related to a lesser number of participants in Covaxin arm compared to the Covishield or due to the difference in characteristics of participants or related to differential immunogenic response due to the difference in loading dose of antigen in vaccine - is not exactly known - and need further studies. Nevertheless, even in age-, sex- and BMI-matched propensity analysis, responder rate was 2-fold higher with Covishield as compared to the Covaxin recipient. Notably, presence of any comorbidities, past history of SARS-CoV-2 infection and types of vaccine used were an independent predictor of antibody response in multiple regression analysis.

Our findings are both similar and dissimilar to published evidence in randomized controlled trials. In phase 1/2 trial in humans, one dose of ChAdOx1 nCoV-19 (Covishield) vaccine elicited a significant increase in IgG antibodies (peaked by day 28) against SARS-CoV-2 spike protein, as measured by ELISA in 127 participants [11]. Similarly, in phase 2 Covaxin trial, there was a significant increase in IgG anti-spike antibody titre at day 28 (on the day of second dose), measured by ELISA in 380 participants (190 participants each in 3 mcg and 6 mcg dose of Algel-IDMG Covaxin). Notably, seroconversion rate (defined as post-vaccination IgG anti-spike antibody titre 4-fold higher than the baseline) have varied from 65.0% to 71.2% at day 28 with 6 mcg and 3 mcg Algel-IMDG dose of Covaxin, respectively [13]. Nevertheless, results from our cross-sectional study are similar in line with other real-world studies that have measured anti-spike antibody response following first dose of other approved vaccines, although none correlated it with comorbidities. In a cross-sectional community survey from England (REal-time Assessment of Community Transmission-2 program, REACT-2) that studied IgG anti-spike antibody kinetics after a single dose of Pfizer-BioNTech vaccine involving 3,011 participants showed 84.1% seropositivity (unadjusted) in people under 60 years of age, tested at 21 days or after but before the second dose [15]. This IgG seropositivity increased to overall 91.1% after the booster dose across all age groups. There was a consistent decreasing trend in IgG positivity with the increasing age in REACT-2 study. The prevalence of IgG positivity rate at or after 21 days after the first dose of Pfizer-BioNTech vaccine was 94.7% in age group of 18-29 years which was found to be reduced to 34.7% in age ≥ 80 years, although this prevalence rate increased to 100.0% and 87.8% in both age group, respectively 21 days or more after the booster dose. A high IgG positivity of 90.1% to a single dose of Pfizer-BioNTech vaccine was observed in people with past confirmed or suspected COVID across all age groups. Antibody response evaluated gender-wise in REACT-2 study also suggested that at any point of time binding antibody titre was consistently higher in females compared to the males after the first dose, tested at 21 days or more. Another study of UK healthcare worker also suggested an inverse correlation between age and anti-spike antibody response (significantly higher anti-spike antibody in age <50 years vs. ≥ 50 years) following a single dose of Pfizer-BioNTech vaccine (Prendecki 2021, in press) [15]. A study from Israel involving 514 health care workers that studied the anti-spike antibody response at day 21 or after but before the second dose following the first dose of Pfizer-BioNTech vaccine using the same kit we used (LIASON^®^ S1/S2 quantitative antibody detection kit, DiaSorin Saluggia, Italy), reported a 92.0% (475/514) responder rate [16]. The geometric mean concentration (GMC) in responders was 68.6 AU/ml (95% CI, 64.0 - 73.6 AU/ml). However, a significant decrease in responder rate was observed with the increasing age. Amongst the 7.6% (39/514) non-responders, older age (54 vs. 45 year; p <0.001) and Jewish ethnicity (p=0.01) significantly represented the most. Although no significant difference in antibody responders was observed gender-wise, higher trend of anti-spike antibody titre GMC was observed in females (75.9 AU/ml) compared to the males (64.6 AU/ml). Notably, none of these studies reported the antibody response in relation to comorbidities. A recent pre-print study has reported a significantly diminished anti-spike antibody titre in 81 hemodialysis patients compared to 80 healthy control even after the two completed doses of Pfizer-BioNTech vaccine [17].

Nonetheless, the moot question is - does humoral antibody response to a vaccine correlates with the efficacy (reduction in severity and mortality due to COVID-19, acquired after vaccination)? Although no such direct studies are currently available and correlation of antibody titre to the vaccine efficacy is less well understood till date, neutralizing antibody targeting different epitopes of spike glycoprotein have been demonstrated to protect from COVID-19 with ChAdOx1 nCoV-19 (Covishield) and Moderna mRNA-1273 vaccine in pre-clinical studies [18, 19]. Moreover, neutralizing antibody titre following vaccination was highly correlated with the neutralizing antibody in convalescent post-SARS-CoV-2 infection. Available evidence with Covaxin has also demonstrated a strong immunogenic response in pre-clinical studies [20]. Notably, phase 1/2 clinical trials have also demonstrated a favorable T-cell response with Covishield up to 8-weeks after a single dose of Covishield [21]. Despite these findings it is not exactly known as to what level of binding and neutralizing antibody protects human from COVID-19 [22]. Interestingly, at least one longitudinal study found no relationship between post-vaccination serum binding-antibody in SARS CoV2 naїve individuals suggesting assessment of short-term antibody titers alone may fail to predict long-term immunity conferred by the vaccines [23].

To best of our knowledge, this large Pan-India cross-sectional study would be the first of its kind that involved participants (all doctors) from 13 States and 22 cities, and reported anti-spike antibody kinetics after the first dose of two different vaccines. However, we also acknowledge several limitations. Firstly, an important consideration for a cross-sectional study design lies in the randomly selected sample obtained from the targeted population for which the results would be generalized. However, in the present study, we have used a convenience sampling amounting to selection bias. Moreover, to answer a research question like antibody response rate, a community-based study in a larger population with multi-stage sampling would have be an ideal sampling method. Furthermore, for the outcome like responder rate, we could not use stratification by the age and sex. Secondly, we used a binary logistic regression (to identify the predictors of non-response to vaccines) which primarily assumes linearity between the explanatory variable and the outcome variable, hence this model may miss out any predictor variable which may have non-linear relationship with the outcome variable. Thirdly, we have measured only anti-spike binding antibody and could not assess neutralizing antibody as well as cell-mediated immune response such as Th-1 and Th-2 dependent antibody or cytokines (primarily due to the lack of standardized commercial labs in India). Furthermore, we could not measure the baseline anti-spike antibody titre prior to the vaccination, because of delay in finalizing the logistic of this study. Finally, a single value of short-term anti-spike antibody as reported in this preliminary report may not necessarily predict the efficacy of vaccine, nor the absence of seropositivity confer failure of vaccine after the first dose. Therefore, a serial evaluation of antibody kinetics especially the memory B cell response after the completion of second dose and subsequent longer assessment would be an ideal reflection of humoral response following vaccination.

In conclusion, this preliminary report of a cross-sectional study suggests that in age, sex and BMI matched propensity analysis the first dose of Covishield induces significant binding-antibody immunoreactivity by increasing anti-spike antibody in 88.4% individuals, day 21 onwards after the first dose. In contrast, Covaxin showed positive immunoreactivity in 43.0% of participants after the first dose. This suggest that 11.6% and 57.0% participants in Covishield and Covaxin arm respectively, do not show adequate rise in anti-spike antibody after the first dose. Age >60 years and presence of any comorbidities including HTN and T2DM appear to reduce the response rate after the first dose of vaccines. Ongoing COVAT study will further advance our knowledge on anti-spike antibody kinetics after the second dose of two vaccines until 6-month post-vaccination.

## Data Availability

All the authors are responsible for the originality of this study. Original data can be shared from first author if necessary, after a reasonable request.

## Acknowledgments

We would like to thank all the participants who volunteered for this study. We express our sincere gratitude and acknowledgment to our Indian regional coordinators for the smooth conduct of this study that include (in alphabetical order) – Drs. Akash Kumar Singh (Vadodara), Amit Gupta (Noida), Anuj Maheshwari (Lucknow), Arvind Kumar Ojha (Kolkata), Bhavtharini (Erode), B. Harish Kumar (Mysore), J K Sharma (New Delhi), Jayant Panda (Cuttack), Kavyachand Yalamudi (Guntur), Kiran Shah (Vadodara), M Gowri Sankar (Coimbatore), Manohar KN (Bangalore), Meena Chhabra (New Delhi), Pratap Jethwani (Rajkot), M Shunmugavelu (Trichy), Rajiv Kovil (Mumbai), Sunil Gupta (Nagpur), Subhash Kumar (Patna), Somnath (Hyderabad), Urman Dhruv (Ahmedabad). Our heartfelt thanks to Ms. Roma Dave (Dietician) and Dr. Priya Phatak (Ahmedabad) for keeping entire data up-to-date and confidential at every step.

## Contribution of authors

AKS and SRP conceptualized and designed the study. NKS, AG and AS monitored the study and captured the data at all point of time. AKS and KB conducted the statistical analysis. AKS and RS wrote the first draft. AKS, KB and RS revised the manuscript. All authors gave their intellectual inputs while preparing the manuscript and agreed mutually to submit to this journal.

