## Supplemental Table 1 for "Antibody Response after First-dose of ChAdOx1-nCOV (Covishield™^®^) and BBV-152 (Covaxin™^®^) amongst Health Care Workers in India: Preliminary Results of Cross-sectional Coronavirus Vaccine-induced Antibody Titre (COVAT) study"

**Supplementary table 1:** **Antibody titer (Median, IQR) between Day 21-28, 29-36 and 36-45 days after the first dose of each vaccine**

|  | Day, n | Antibody Titre  Median (IQR) | P |
| --- | --- | --- | --- |
| Overall, N= 552 | Day 21-28, n=378  Day 29-36, n = 128  Day 37-45, n = 46 | 57 (21-122)  42 (18-102)  63.5 (40.75-100.75) | 0.260 |
| Covishield, N= 456 | Day 21-28, n=305  Day 29-36, n = 106  Day 37-45, n = 45 | 63 (30-155)  55.5 (25.25-107)  64 (44-101.5) | 0.418 |
| Covaxin, N= 96 | Day 21-28, n=73  Day 29-36, n = 22  Day 37-45, n = 01 | 6 (4-103)  7 (4-83.25)  4 (4-4) | 0.603 |
